# Breaking down causes, consequences, and mediating effects of age-related telomere shortening on human health

**DOI:** 10.1101/2024.01.12.24301196

**Authors:** Samuel Moix, Marie Sadler, Zoltán Kutalik, Chiara Auwerx

## Abstract

Telomeres represent repeated DNA sequences at the ends of chromosomes, which shorten with each cell division. Factors modulating telomere attrition and the health consequences thereof are not fully understood. To address this, we leveraged data from 326,363 unrelated UK Biobank participants of European ancestry and used linear regression and bidirectional univariable and multivariable Mendelian randomization (MR and MVMR) to elucidate the relationships between leukocyte telomere length (LTL) and 141 complex traits, including diseases, biomarkers, and lifestyle factors. We confirm that telomeres shorten with age and show a stronger decline in males than in females, which cannot be explained by hormonal or lifestyle differences. MR revealed 23 traits modulating LTL; e.g., smoking cessation and high educational attainment associated with longer LTL, while weekly alcohol intake, body mass index, urate levels, and female reproductive events, such as childbirth, associated with shorter LTL. We also identified 26 traits affected by LTL, with risk for cardiovascular, pulmonary, renal, and some autoimmune diseases being increased by short LTL, while longer LTL increased risk for other autoimmune conditions and cancers. Through MVMR, we show that LTL partially mediates the impact of educational attainment, body mass index, and female age at childbirth on lifespan. These results provide new insights into the biology of telomere regulation by shedding light on the modulators, consequences, and the mediatory role of telomere shortening, portraying an intricate relationship between LTL, diseases, lifestyle, and socio-economic factors.

## Introduction

Aging represents a leading risk factor for diseases such as cancer, cardiovascular diseases, and neurodegeneration [1]. Chronological age fails to account for individual differences in aging rates and vulnerability to diseases [2]. Biological age intends to address this limitation by reflecting the physiological state of an individual and accounting for variations in cellular and tissue health. Several biomarkers can be used to estimate biological age, with DNA methylation being particularly popular due to its availability across tissues, and its sensitivity to both disease states and environmental factors [3, 4, 5]. However, given the complex nature of the aging process, additional biomarkers beyond DNA methylation are required to fully understand the underlying causes and mechanisms of aging and age-related diseases [6].

One such biomarker is telomere length. Telomeres are DNA repeats at chromosome ends that act as protective caps against genomic degradation. As organisms age, they undergo an increasing number of cell divisions, leading to an incremental decrease in telomere length. Acting as mitotic clocks, telomeres shorten until they reach a critical length, triggering cellular senescence and/or apoptosis [7]. Consequently, shorter telomeres have been associated with lifestyle factors, including smoking [8], reduced physical activity [9], high processed meat and low fruit consumption [10, 11], as well as a wide range of diseases, from pulmonary [12], renal [13], and metabolic [14] disorders to cancer [15, 16]. Paradoxically, longer telomeres have also been associated with poor health outcomes, especially cancers [17]. However, most studies so far were limited in the number of studied traits, relied on small sample sizes, and did not probe the directionality of the established associations.

Recently, efforts to assess leukocyte telomere length (LTL) in large population biobanks have allowed comprehensive exploration of its relationships with lifestyle factors and health outcomes. Performing an LTL phenome-wide association study in 62,271 participants from the Vanderbilt University and Marshfield Clinic biobanks, Allaire *et al*., identified associations with 67 phenotypes and showed that both shorter and longer telomeres associated with increased mortality [18]. Release of LTL measurements for ∼500,000 UK Biobank (UKBB) participants [19] and the companion first large-scale telomere length genome-wide association study (GWAS) [20] prompted investigation of the impact of LTL on hundreds of traits using phenome-wide Mendelian randomization (MR) [20, 21]. These studies showed that longer LTL increases risk for neoplastic and genitourinary diseases while lowering risk for respiratory, digestive, and cardiovascular disorders [20, 21], with about 40% of these associations confirmed when using FinnGen disease association summary statistics [21].

Our study builds on this body of work by dissecting observational correlations between LTL and 141 traits into causes and consequences through a bidirectional MR causal framework (Figure 1). Additionally, we performed sex-stratified analyses and used multivariable Mendelian Randomization (MVMR) to disentangle the interplay between LTL and various traits, with a particular focus on the mediating role of LTL in longevity. Together, we identify traits influencing LTL, and how in turn the latter impacts the human phenome, contributing to a deeper understanding of telomere biology and its relation to health.

**Figure 1.**
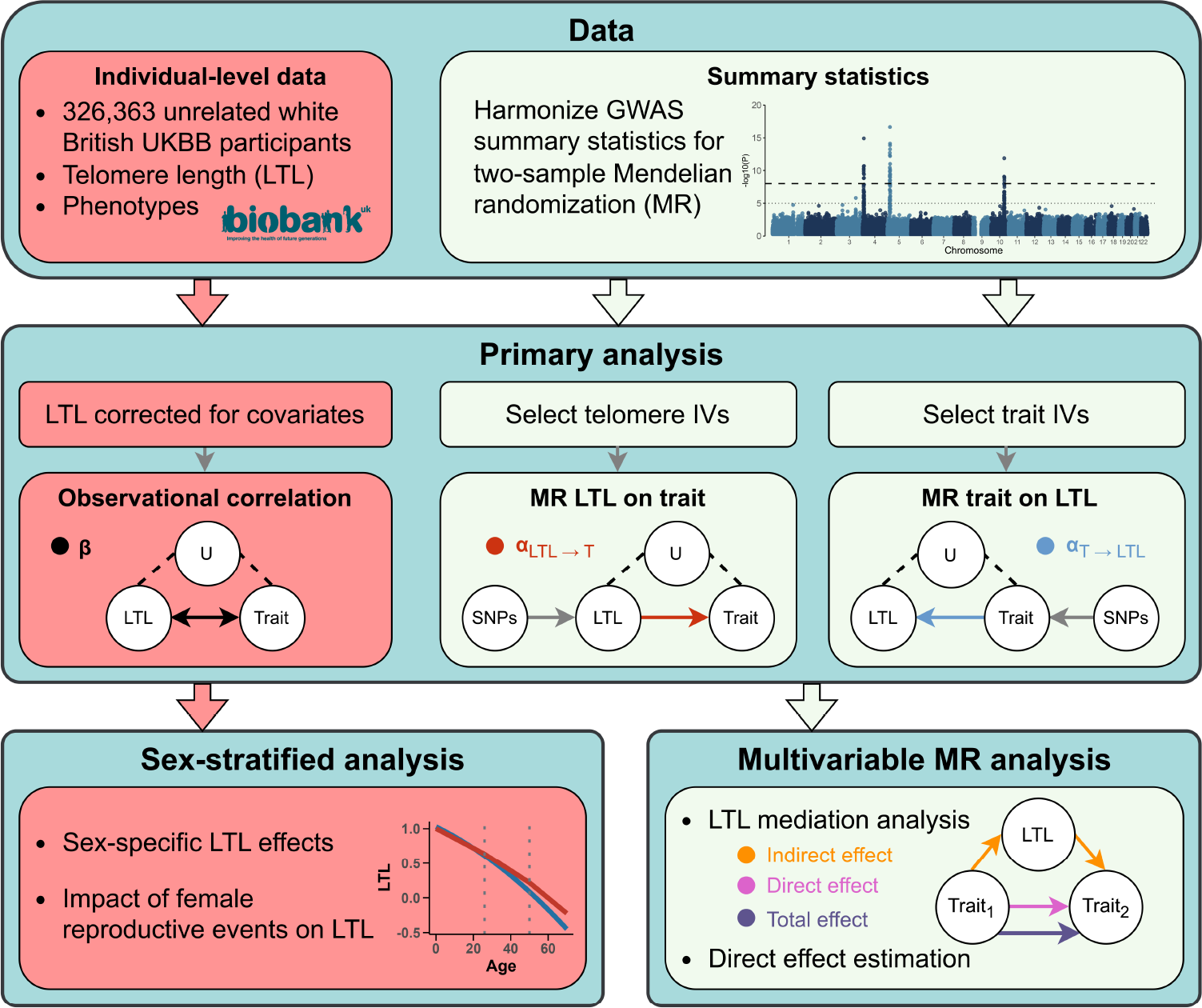
Schematic representation of the study’s workflow. Red and light green boxes denote steps using individual-level phenotypic data from the UK Biobank and GWAS summary statistics, respectively. Top: Data extraction process. Middle: Analyses focused on LTL trait relationships including observational correlation (*β*; black), Mendelian randomization to assess the impact of LTL on traits (*α*_*LT L*→*T*_ ; red), and Mendelian randomization to assess the impact of traits on LTL (*α*_*T* →*LT L*_; blue). LTL covariates comprise age, age^2^, array, sex, and the interaction of the latter with the priors. U = unmeasured confounding factors; IVs = instrumental variables (i.e., genetic variants with genome-wide significant association to the considered trait). SNP = single nucleotide polymorphism. Bottom: Follow-up analyses including sex-specific LTL effects and LTL mediation analysis through multivariable Mendelian randomization.

## Materials and methods

### Data

#### Individual-level UK Biobank data

Observational analyses were carried out in the UK Biobank (UKBB), a cohort of ∼500,000 volunteers from the general UK population aged between 40–69 years at recruitment [22]. Phenotype data were accessed through approved application 16389. Analyses were conducted on 326,363 participants with known sex, age, and LTL after the exclusion of individuals of non-white and non-British ancestry (self-reported + genetically defined), relatives (≤ 3^*rd*^ degree), and gender mismatches (see UKBB Resource 531), as well as those who retracted their participation. Given that LTL measurements are derived from blood, we further excluded 4,376 individuals with blood malignancies, based on self-reports (UKBB field #20001 codes 1047, 1048, 1050, 1051, 1052, 1053, 1055, 1056, 1058) or hospital diagnoses (#41270; International Classification of Diseases 10th Revision [ICD10] codes mapping to the PheCode “cancer of lymphatic and hematopoietic tissue” [23]).

We used technically adjusted and standardized LTL (#22192) [19] and assessed its relation to 166 complex traits (Table S1). These include 60 common diseases defined based on hospital diagnoses (#41270; last diagnosis September 2021), while excluding from controls individuals with self-reported (#20001, #20002) or hospital-diagnosed (#41270) conditions related to the investigated disease [24]. Disease phenotypes were used to calculate a disease burden phenotype, i.e., the total number of diseases diagnosed in an individual among the 60 considered ones. The remaining 105 traits include 11 anthropometric traits (e.g., weight), 41 biomarkers (e.g., serum lipids), 18 life events (e.g., age at menarche and menopause), 26 lifestyles (e.g., beef intake) and socio-economic factors (e.g., Townsend deprivation index), and 9 miscellaneous traits. Definitions of composite phenotypes are described in the Supplementary Note. Briefly, continuous traits with multiple instances were averaged, while the first instance was used for integers or factors. To minimize noise, outliers (mean ± 5 standard deviations [SD]) in continuous traits were removed. Factorial variables were numerically converted for efficient integration into the regression model. All traits, including binary predictors, were then scaled to have zero mean and unit variance to obtain more comparable effect sizes. As the 167 assessed traits (i.e., 166 above-mentioned + blood cancer) were partially correlated we estimated the number of effective tests [25], i.e., the number of tests needed to explain 99.5% of the variance in our phenotypic dataset, to 141, resulting in a significance threshold of *p* < 0.05/141 = 3.5*e*−4 for observational correlation and MR analyses.

#### GWAS summary statistics

When available (i.e., for non-composite traits), genome-wide association study (GWAS) summary statistics originate from the Neale group (file release July 2018; http://www.nealelab.is/uk-biobank) (Table S2). Summary statistics for reproductive lifespan were derived from GWAS on age at menopause and menarche by first backtransforming the effects on year-scale and then computing their difference:

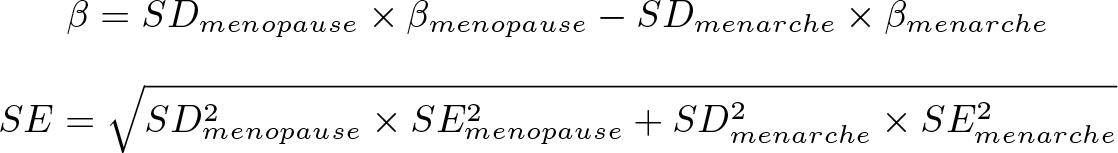

The sample size for the resulting summary statistic was set to the lowest of the two (i.e., age at menopause; *N* = 111, 593) and p-values were computed with a two-sided test based on a t-statistic obtained by dividing the effect size by its standard error. For diseases, a set of previously compiled GWAS summary statistics [26] of predominantly European-descent consortia meta-analyses was used (Table S2). Summary statistics were harmonized with the UK10K reference panel [27] and restricted to autosomal chromosomes. After excluding palindromic single-nucleotide polymorphisms (SNP) and adjusting strand-flipped SNPs, effect sizes were standardized to represent the square root of the explained variance [28].

### Observational correlation

#### Predictors of LTL variability

To estimate the fraction of LTL variability explained by the human phenome, we used Lasso regression (glmnet package in R [29]) with unadjusted normalized LTL as the outcome variable and traits with less than 5%, 7%, and 10% missing data as possible predictors in a joint model. Given the non-deterministic choice of the optimized regularisation parameter (one SE rule lambda), 50 regressions were fitted and traits that were selected in at least 95% of the cases were considered as predictors.

#### Single trait linear regression

We adjusted LTL for age, age^2^, genotyping array, sex, and the interaction of the latter with the priors and used this variable as the outcome in 166 linear regression models with the traits described in Table S1 as explanatory variables. Effect sizes reported in text are in SD_LTL_/SD_Trait_, except for the effect of age, in which case effects are reported in SD_LTL_/year. We followed up on specific associations with sensitivity analyses to identify possible confounders:

- In individuals using cholesterol-lowering drugs (#6177 and #6153), serum lipids levels were corrected for the average simvastatin effect, i.e., + 1.6 mmol/L, 1.4 mmol/L, 0.4 mmol/L, −0.1 mmol/L of total cholesterol, low-density lipoprotein (LDL), triglycerides and high-density lipoprotein (HDL), respectively [30].
- Reproductive traits showing significant (*p <* 0.05/141) association with LTL were corrected for socioeconomic status (SES; i.e., Townsend deprivation index (TDI; #189), average total household income before tax (#738), and educational attainment (EA; see Supplementary Note)).
- In addition to age, sex, and array, LTL was corrected for eosinophil (#30150), lymphocyte (#30120), monocyte (#30130), neutrophil (#30140), platelet (#30080), red blood cell (#30010), reticulocyte (#30250), and white blood cell (#30000) counts and linear regressions with non-blood trait count traits were performed anew to ensure the LTL associations were unbiased. As a result, the available sample size reduced to *N* = 308, 346.

#### Sex-stratified analysis

Sex-stratified linear regression of LTL on age was assessed, as well as of LTL corrected for age, age^2^, and genotyping array on non-sex-specific traits. Sex-specific differences were identified by comparing male and female effects and deemed strictly significant at *p <* 0.05/141 and nominally significant at *p <* 0.05 (see Effect size comparison). We investigated the possible impact of sex hormone levels (i.e., sex hormone-binding globulin (SHBG) and testosterone), as well as lifestyle factors (i.e., fruit (#1309), vegetable (#1289), beef (#1369), alcohol intake frequency (#1558), smoking status (#20116) and weekly alcohol intake (see Supplementary Note)) on sex differences in age-dependent telomere shortening by controlling for these factors in our regression models.

#### Female reproductive phases

To assess the impact of childbearing and menopause on LTL, we identified three distinct female reproductive phases: (1) years before first childbirth, (2) premenopausal years after first childbirth, and (3) postmenopausal years. Number of years spent in each phase was derived from current age (#21003), age at first childbirth (#2754), and age at menopause onset (#3581). Phases (2) and (3) were set to 0 for females with no children (#2734: number of births = 0) and premenopausal women, respectively. The joint linear regression model included time spent in each phase and two indicator variables for whether the women carried a pregnancy to term and experienced menopause. Female participants who had their first child post-menopause, lacked a menopausal status (#2724) or age at menopause (#3581), or did not specify childbirth events (#2734) or age at first childbirth (#2754) were excluded from this analysis.

## Mendelian randomization

### Bidirectional univariable Mendelian randomization

GWAS summary statistics were used to conduct bidirectional two-sample Mendelian randomization (MR), with *α*_*LTL*→*T*_ representing the causal impact of LTL (exposure) on complex traits (outcome) and *α*_*T* →*LTL*_ the causal impact of complex traits (exposure) on LTL (outcome) (Figure 1). Harmonized SNPs significantly associated (*p* < 5*e*−8) with the exposure were clumped (*p*1 = 0.0001, *p*2 = 0.01, *kb* = 250, and *r*2 = 0.01) with PLINK v1.9 [31] and retained as instrumental variables (IVs). As the *HBB* gene was used as a control for the LTL measurements [20], we removed the single SNP (rs1609812, *p* = 3.9*e*−65) associated with this gene (chr11:5*’*246*’*696-5*’*248’301; GRCh37/hg19) to prevent spurious associations. Due to the complex long-range linkage disequilibrium (LD) structure of the HLA locus, 30 SNPs found on the extended HLA region (chr6:25’000’000–37’000’000; GRCh37/hg19) were also removed from our IVs [32]. Further IVs were removed based on exposure-outcome harmonization, difference in allele frequency (≥ 0.05), and Steiger filtering (*Z* ≤ −1.96). Bidirectional MR analyses were carried out with the TwoSampleMR R package (v0.5.6) [33], primarily through the inverse variance weighted (IVW) method. LTL on trait and trait on LTL MR effects were computed for 151 and 141 traits, respectively, with at least two IVs.

Sensitivity analyses were conducted using additional MR methods, i.e., MR Egger, simple mode, weighted median, and weighted mode, to ensure robustness of the results. LTL on trait and trait on LTL MR analyses with these methods were carried out for 151 and 134 traits with at least three IVs, respectively. Heterogeneity was assessed using Cochran’s Q-statistics. Given a high proportion of elevated Q-statistics, we additionally run MR-PRESSO [34] for relationships with significant IVW MR effects. To further ensure that our results are not biased by pleiotropy - which violates the MR assumption that IVs only affect the outcome through the exposure [35] - we first filtered genome-wide significant exposure SNPs and harmonized these SNPs across all 152 traits with available GWAS summary statistics (i.e., 151 traits + LTL). Harmonized GWAS data were clumped. We next applied Steiger filtering between the exposure and all other traits to ensure that the selected SNPs are more strongly associated with the exposure than with any of the other included traits. SNPs that passed filtering for all traits were retained as IVs and MR analyses were conducted on these. This approach serves as a reasonable pleiotropy filter due to the diverse nature of our phenotypes.

#### LTL mediation analysis

Excluding hematological traits due to potential confounding, we used multivariable MR (MVMR) to assess the mediating role of LTL between 18 LTL-associated traits (*p* < 0.05/141) and lifespan (proxied from parental lifespan [36]). We further examined the global mediatory role of LTL between these 18 LTL-affecting traits and 19 traits causally impacted by LTL (*p* < 0.05/141). This corresponded to 359 pairs (18 * 20 (i.e., 19 traits + lifespan), excluding one pair as insulin-like growth factor 1 (IGF-1) associated with LTL as both exposure and outcome), setting the significance threshold for the total and indirect effects at *p* < 0.05/359 = 1.4*e*−4. During SNP harmonization, exposure IVs (i.e., trait IVs) were prioritized over mediator IVs (i.e., LTL IVs). Steiger filtering was applied to both exposure IVs with respect to outcome and mediator and to mediator IVs in relation to the outcome. Indirect effects were determined through two strategies: difference in coefficients and product of coefficients [37]. The former subtracts the direct effect (MVMR) from the total effect (IVW), while the latter multiplies the univariable MR estimates from the exposure on the mediator by the MVMR effect of the mediator on the outcome. Both approaches generated consistent results and we present the product of coefficients method due to easier interpretability in the main text. We further corrected these estimates for regression dilution bias [28]. Mediation proportions (*P*_M_) represent the ratio of the indirect (*α*_*indirect*_) to total (*α*_*total*_) effect with 95% confidence intervals (upper limit capped at 100%) estimated from the 2.5th and 97.5th quantiles of the distribution of 10,000 simulated ratios drawn from 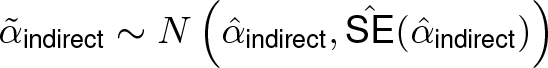 and 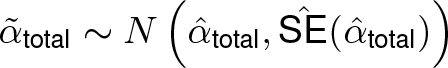.

#### Multi-trait analysis for direct effect estimation

For MVMR with multiple exposures and no predefined mediator, IVs were selected through a two-step process [38]. First, SNPs for each exposure were ranked according to their p-values (more significant p-values receiving lower ranks) and minimum rank across all exposures was determined for each SNP. This minimum rank was used to prioritize SNPs in a subsequent clumping process. IVs were filtered as previously described. Finally, MVMR regression estimates were compared to univariable MR estimates (see Effect size comparison). For the univariable MR, we either used the same IVs as in the MVMR or employed a subset of IVs, which were retained after Steiger filtering between both the outcome and the exposure of interest, as well as between the exposure of interest and the other exposures. We report weak instrument bias via conditional F-statistics [39] and heterogeneity through Cochran’s Q-statistic [40] (see Table S3).

### Effect size comparison

Significant differences between two estimated effect sizes 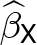 and 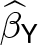 were assessed with a two-sided p-value (*p*_diff._) derived from:

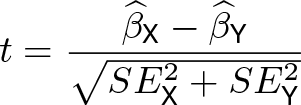

which assumes that the two estimates are uncorrelated. Often these estimates have a positive correlation (as estimated from the same data) and hence the *t*-statistic has a variance smaller than one, thus the test is conservative. This approach was used throughout the study to assess the effect of sensitivity analyses, compare sex-specific regression estimates, and compare univariable MR and MVMR results.

## Results

### Age and sex are the main predictors of LTL variability

Consistent with previous research [41], LTL significantly associated with both age (*β* = −0.023; *p* < 2.2*e*−308) and sex (*β* = 0.091; *p* < 2.2*e*−308), with a stronger (*p*_diff._ = 1.4*e*−25) decline in males (*β*_*males*_ = −0.025) than in females (*β*_*females*_ = −0.021) (Figure S1). To further explore factors contributing to LTL variability, we included 80 traits (Table S1) with *<* 7% missingness rates as predictor variables in a Lasso regression model. Traits retained included age, sex, educational attainment (EA), waist-to-hip ratio (WHR), insulin-like growth factor 1 (IGF-1), urate, and cystatin C levels, along with four blood parameters (Table 1). Among these, LTL was found to be positively associated with female sex, higher EA, and higher IGF-1 levels, while it negatively correlated with the remaining traits. Age and sex accounted for 4.33% of the observed variance in LTL. Incorporating the nine additional above-mentioned traits increased the explained variance to 5.39%. Repeating the analysis with missingness rate thresholds of 5% and 10% retained twelve and seven traits in addition to age and sex, which together explain 5.42% and 5.36% of variability in LTL, respectively, confirming the limited predictive power of the phenome over LTL variability.

**Table 1.** Main determinants of leukocyte telomere length. Effect sizes with 95% confidence intervals from a joint linear regression model for traits with less than 7% missing data retained as significant LTL predictors by lasso analysis. EA = educational attainment; IGF-1 = insulin-like growth factor 1; WHR = waist-to-hip ratio; MCH = mean corpuscular hemoglobin.

| Trait | Estimate | Lower 95% CI | Upper 95% CI | P-value |
| --- | --- | --- | --- | --- |
| Female | 0.071 | 0.066 | 0.077 | $2.33 \times 10^{-160}$ |
| EA | 0.042 | 0.038 | 0.046 | $1.95 \times 10^{-111}$ |
| IGF-1 | 0.028 | 0.024 | 0.032 | $3.57 \times 10^{-49}$ |
| WHR | -0.002 | -0.007 | 0.004 | $5.43 \times 10^{-1}$ |
| Urate | -0.009 | -0.014 | -0.004 | $1.16 \times 10^{-4}$ |
| Monocyte count | -0.017 | -0.021 | -0.013 | $4.89 \times 10^{-17}$ |
| Eosinophil count | -0.021 | -0.025 | -0.017 | $1.00 \times 10^{-28}$ |
| Cystatin C | -0.026 | -0.030 | -0.022 | $4.93 \times 10^{-33}$ |
| Lymphocyte count | -0.039 | -0.043 | -0.035 | $7.43 \times 10^{-87}$ |
| MCH | -0.058 | -0.062 | -0.055 | $4.98 \times 10^{-223}$ |
| Age | -0.155 | -0.159 | -0.151 | $< 2.2 \times 10^{-308}$ |

### LTL broadly associates with complex traits

Age and sex being the major determinants of LTL, we adjusted LTL for age, age^2^, genotyping array, sex, and the interaction of the latter with the priors and regressed adjusted LTL on 166 traits through linear regression, identifying 100 significant associations (*p* < 0.05/141; Table S4). We observed a negative association between the disease burden and LTL (*β* = −0.027; *p* = 1.2*e*−52), suggesting that LTL acts as a global health indicator. The largest effect sizes were noted for father’s (*β* = 0.094; *p* = 4.4*e*−144) and mother’s (*β* = 0.088; *p* = 8.5*e*−216) age at birth, which positively associated with LTL (Figure 2a). In jointly modeling LTL as a function of both parental ages at birth and the participant’s age, sex, their interaction, and EA, we found that the observed associations were independent of the participant’s education level (Figure S2). Participant’s education level likely echoes parental EA [42] and indirectly affects parental age at birth, indicating that the association is likely not confounded by socio-economic factors and genuinely driven by older parental age at birth. Next, for the 141 traits with available GWAS summary statistics and at least two IVs, we inferred bidirectional causal relationships through univariable MR, identifying 23 significant causal effects of traits on LTL (*α*_*T* →*LTL*_) and 26 significant effects of LTL on traits (*α*_*LTL*→*T*_) (*p* < 0.05/141; Figure 2; Table S4). As a sensitivity analysis, we re-analyzed IVW-significant relations with MR Egger, simple mode, weighted median, weighted mode, and MR-PRESSO (Figures S3-4). Furthermore, we ran IVW MR with a restricted set of IVs stringently selected to minimize the pleiotropy assumption violation (see Methods; Figure S5).

**Figure 2.**
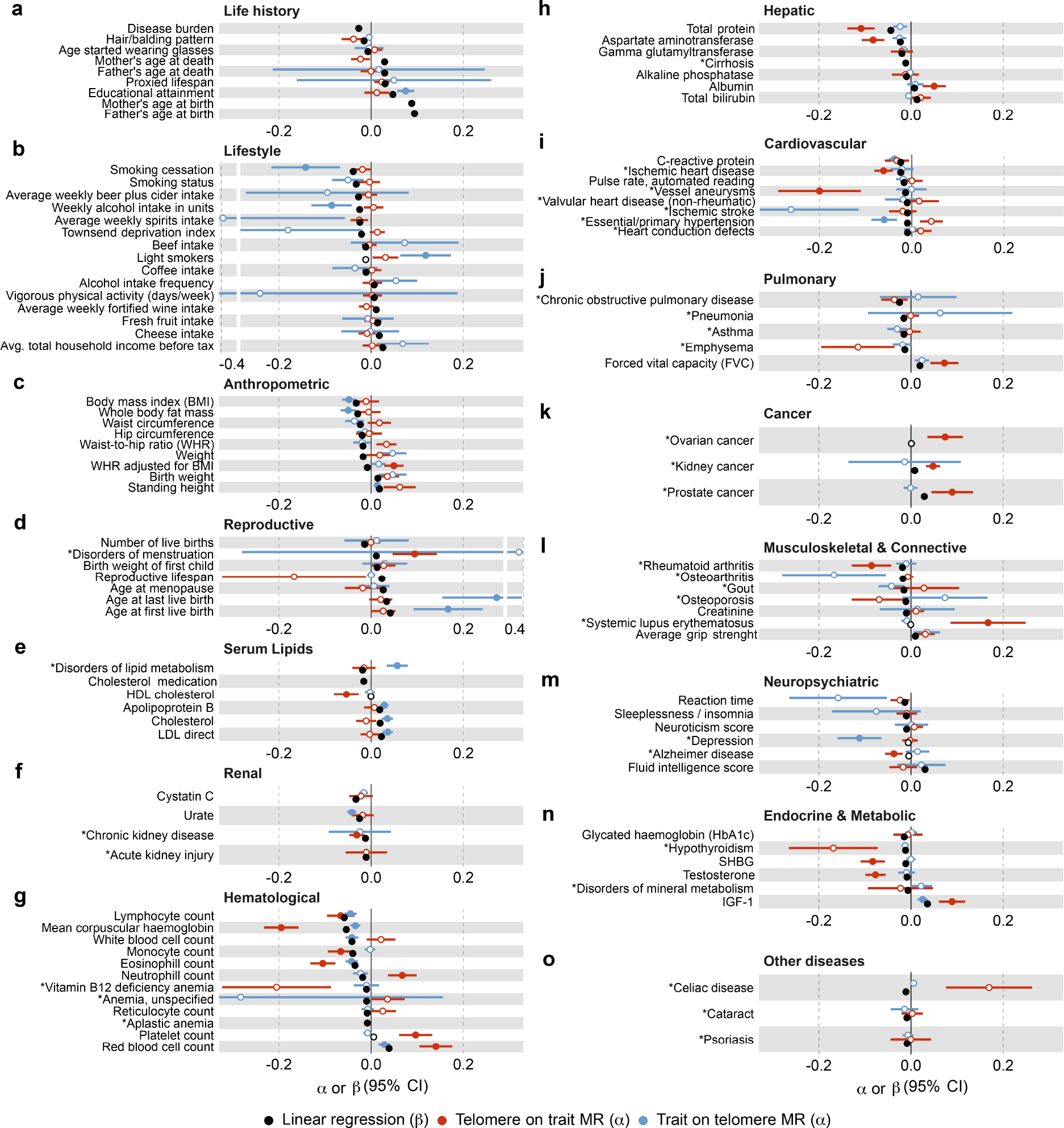
Observational and causal associations between traits and LTL. Estimates (x-axis) with 95% confidence intervals (CI) for traits (y-axis) with at least one strictly significant (*p* < 0.05/141) association with LTL across the observational correlation (linear regression; *β*; black) and inverse weighted-variance (IVW) Mendelian randomization (MR) estimates of LTL on trait (*α*; red) and trait on LTL (*α*; blue) are shown. Strictly significant effects are shown as full circles; otherwise as empty circles. For diseases (*), one SD change in LTL corresponds to one log(*OR*) change, implying a scale of SD_LTL_/log(*OR*) for the effects of diseases on LTL, and log(*OR*)/SD_LTL_ for the effect of LTL on the disease, so that observational effects and MR effects are not directly comparable (see Table S4).

### Modulators of LTL

#### Lifestyle and environmental factors

Our results are overall concordant with deleterious lifestyle habits leading to shorter LTL (Figure 2b). A negative correlation was observed between smoking cessation and LTL (*β* = −0.039; *p* = 9.4*e*−50), mirrored by a detrimental causal effect of failure to quit smoking on LTL (*α*_*T* →*LTL*_ = −0.142; *p* = 1.8*e*−4). Alcohol consumption, measured as total weekly intake of alcohol units, also exhibited a negative causal effect on LTL (*α*_*T* →*LTL*_ = −0.086; *p* = 1.3*e*−4), while beef consumption showed a mere associative (*β* = −0.012; *p* = 2.4*e*−11) but no causal link (*p* = 0.223). Conversely, healthy habits such as high fresh fruit intake (*β* = 0.014; *p* = 6.4*e*−15) and physical activity (*β* = 0.007; *p* = 1.7*e*−4) displayed positive associations with LTL, as did SES captured by average household income (*β* = 0.025; *p* = 1.1*e*−40) or EA (*β* = 0.047; *p* = 1.9*e*−155), even though only the latter showed clear causal evidence (*α*_*T* →*LTL*_ = 0.075; *p* = 2.2*e*−15). Our data also suggest that the psychological state of an individual can impact LTL as depression causes shorter LTL (*α*_*T* →*LTL*_ = −0.112; *p* = 4.4*e*−6). One possible explanation for this observation is that depression promotes oxidative stress and inflammation, both of which are critical modulators of LTL [7, 43, 44]. The latter is supported by a negative causal effect of the inflammation marker C-reactive Protein (CRP) on LTL (*α*_*T* →*LTL*_ = −0.037; *p* = 9.3*e*−10). Overall, these results highlight the significant impact of lifestyle and environmental factors on LTL and support the paradigm that exposures typically considered as deleterious lead to shorter LTL.

#### Anthropometric traits

We detect several associations with anthropometric traits (Figure 2c). Body metrics such as body mass index (BMI) and body fat mass (BFM) demonstrated significant negative observational correlation (BMI: *β* = −0.032; *p* = 2.4*e*−75; BFM: *β* = −0.029; *p* = 1.2*e*−60) and causal effects on LTL (BMI: *α*_*T* →*LTL*_ = −0.048; *p* = 4.9*e*−10; BFM: *α*_*T* →*LTL*_ = −0.050; *p* = 7.6*e*−9). Conversely, a positive correlation was observed between LTL and height (*β* = 0.018; *p* = 2.2*e*−24), with MR analysis revealing a nominally significant effect of LTL on height (*α*_*LTL*→*T*_ = 0.062; *p* = 4.5*e*−4) and strictly significant effect of height on LTL (*α*_*T* →*LTL*_ = 0.014; *p* = 4.0*e*−5).

#### Female reproductive traits

Observational correlation between LTL and female reproductive traits including age at first (AFB; *β* = 0.042; *p* = 1.4*e*−54) and last (ALB; *β* = 0.034; *p* = 2.5*e*−36) birth, reproductive lifespan (*β* = 0.023; *p* = 3.7*e*−13), age at menopause (*β* = 0.026; *p* = 1.5*e*−16), and menstrual disorders (*β* = 0.011; *p* = 1.7*e*−5) were observed (Figure 2d). Only the effect of AFB (*p*_*diff*._ = 8.0*e*−10) and ALB (*p*_*diff*._ = 0.001) were significantly reduced after accounting for SES, even though they remained significant (Figure S7a). Both traits also causally influenced LTL (AFB: *α*_*T* →*LTL*_ = 0.167; *p* = 1.2*e*−5; ALB: *α*_*T* →*LTL*_ = 0.272; *p* = 6.1*e*−6), suggesting that timing of female reproductive events could modulate LTL. To explore this, we compared LTL in women with and without children, finding shorter LTL in women who had given birth (Welch two-sample t-test: *p* = 7.4*e*−11), suggesting that childbirth could accelerate LTL shortening. We next divided female participants’ age into three reproductive periods: (1) premenopausal before childbirth, (2) premenopausal after childbirth, and (3) postmenopausal, and used the number of years spent in each period as predictors for LTL. LTL shortening accelerated over the course of these periods, with the weakest effect on LTL found for premenopausal years before childbirth (*β* = −0.013; *p* = 3.6*e*−120), followed by premenopausal years after childbirth (*β* = −0.020; *p* = 7.1*e*−233), and postmenopausal years (*β* = −0.024; *p* < 2.2*e*−308) (Figure 3), in line with the hypothesis that female reproductive events trigger acceleration in LTL shortening.

**Figure 3.**
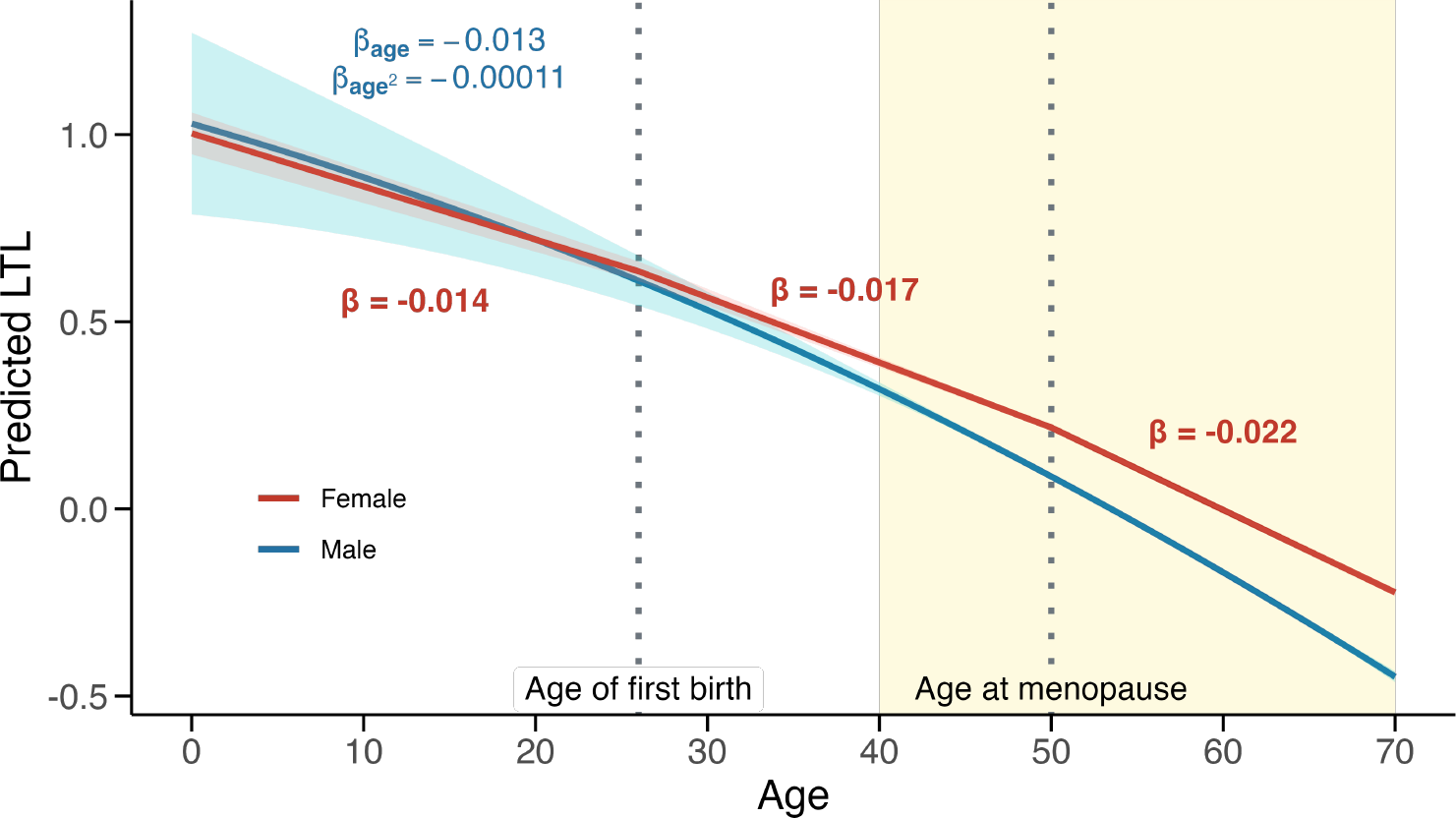
Schematic representation of LTL shortening across different female reproductive life phases. Relation (*β*) between standardized LTL (y-axis) and age (x-axis) across three female reproductive life periods (red). Dotted vertical lines indicate mean age of first birth (26 years) and mean age at menopause (50 years). As a comparison, we depict the quadratic LTL regression in males (*β*_*age*_; *β*_*age*2_ ; blue). 95% confidence intervals are shown for the linear predictions. Yellow background indicates the age range for which data is available (40-70 years) and used to build predictions; regions outside this range are extrapolated for males and estimated from age at first birth and age at menopause information for females.

#### Serum lipids

We found predominantly positive associations between LTL and serum lipid levels, i.e., apolipoprotein B (ApoB; *β* = 0.019; *p* = 9.4*e*−25), total cholesterol (*β* = 0.019; *p* = 2.9*e*−27), and LDL-cholesterol (*β* = 0.022; *p* = 4.2*e*−35) (Figure 2e). After adjusting for cholesterol-lowering drug use, the positive relation between LTL and both total and LDL-cholesterol decreased but remained significant (Figure S7b). LDL-cholesterol (*α*_*T* →*LTL*_ = 0.036; *p* = 4.7*e*−10) and ApoB (*α*_*T* →*LTL*_ = 0.029; *p* = 2.6*e*−10) levels also causally influenced LTL, which was corroborated by weighted mode MR, weighted median MR, and MR-Egger (Figure S3). Consistently, our findings suggest that disorders of lipid metabolism contribute to longer LTL (*α*_*T* →*LTL*_ = 0.057; *p* = 1.4*e*−6), reiterating the association between increased LTL and high serum lipid levels. Due to their correlated nature, MVMR including levels of LDL-cholesterol, ApoB, and triglycerides as exposures could not disentangle their individual contribution to LTL (Figure S6).

#### Renal health

Lasso analysis retained both urate (*β* = −0.025; *p* = 2.1*e*−44) and cystatin C (*β* = −0.033; *p* = 2.2*e*−75) levels as relevant predictors of LTL (Table 1), highlighting the link between kidney function and LTL. As previously reported [45], MR analyses showed that elevated urate levels decreased LTL (*α*_*T* →*LTL*_ = −0.042; *p* = 9.4*e*−18), possibly due to increased cellular stress and reactive oxygen species production [46]. The urate-LTL association was significantly mediated by CRP, confirming the role of inflammation in this process (*P*_M_ = 34.7%; 95%-CI[17.1%; 55.6%]). We also found that shorter LTL increased the risk for chronic kidney disease (CKD; *α*_*LTL*→*T*_ = −0.031; *p* = 1.5*e*−4) (Figure 2f). Urate levels, which are frequently elevated in CKD patients [47], causally affected CKD risk (*α* = 0.034; *p* = 1.5*e*−8) but this link was not mediated by LTL (*P*_M_ = 4.5%; 95%-CI[−8.3%; 18.5%]) (Table S5).

### Consequences of altered LTL

#### Blood cell counts

Hematological traits (e.g., white blood cell count: *β* = −0.042; *p* = 2.9*e*−120; and mean corpuscular hemoglobin (MCH): *β* = −0.054; *p* = 1.7*e*−200) are among the ones showcasing the strongest observational correlation with LTL (Figure 2g). For 4 out of 11 significantly correlated blood traits, we identified bidirectional causal relationships with LTL, with less pronounced effects from traits on LTL (e.g., MCH: *α*_*LTL*→*T*_ = −0.195; *p* = 2.2*e*−24; *α*_*T* →*LTL*_ = −0.034; *p* = 5.2*e*−10). While for MCH, eosinophil, platelet, and red blood cell counts, the effects from LTL on the latter traits were robust across multiple MR methods (Figure S4), only white blood cell count (*α*_*T* →*LTL*_ = −0.042; *p* = 6.7*e*−9) demonstrated a consistent effect on LTL (Figure S3). As telomere length was measured in leukocytes, we cannot exclude that observed associations with blood traits are confounded by other factors, such as blood cell counts. We therefore adjusted LTL for eosinophil, lymphocyte, monocyte, neutrophil, platelet, red blood cell, reticulocyte, and white blood cell counts in addition to core covariates. Regressing this new variable on the same 158 traits (i.e., 166 traits, excluding the 8 blood count traits we corrected for), we obtained highly similar effect sizes (*ρ* = 0.98). Only associations with smoking status (*p*_diff._ = 1.4*e*−9), smoking cessation (*p*_diff._ = 5.7*e*−6), as well as MCH (*p*_diff._ = 3.3*e*−26) were significantly reduced, yet remained significant. Association with total bilirubin (*p*_diff._ = 1.4*e*−5) was lost, while the one with phosphate levels (*p*_diff._ = 8.0*e*−5) became significant (Figure S8).

#### Hepatic biomarkers

LTL associated with levels of the hepatic biomarkers aspartate aminotransferase (AST; *β* = −0.023; *p* = 1.6*e*−37) and albumin (*β* = 0.007; *p* = 7.2*e*−5) (Figure 2h). Accordingly, finding that shorter LTL causally associated with higher AST (*α*_*LTL*→*T*_ = −0.082; *p* = 3.7*e*−11) and lower albumin levels (*α*_*LTL*→*T*_ = 0.050; *p* = 9.1*e*−5), telltales of underlying liver or inflammatory conditions. Overall, these results underscore the potential role of telomere-driven cellular aging in hepatic function deterioration and/or inflammatory processes.

#### Diseases

Short LTL correlated with increased risk for cardiovascular and pulmonary conditions, reflecting previous findings [12, 48]. For instance, LTL had a negative causal impact on aneurysm risk (*α*_*LTL*→*T*_ = −0.200; *p* = 1.2*e*−5) and a positive one on forced vital capacity (*α*_*LTL*→*T*_ = 0.072; *p* = 3.2*e*−6). In line with that, we observed a negative correlation with risk for pulmonary diseases such as emphysema (*β* = −0.013; *p* = 3.5*e*−12) or chronic obstructive pulmonary disease (COPD; *β* = −0.025; *p* = 9.0*e*−40). While the MR effects of LTL on emphysema (*α*_*LTL*→*T*_ = −0.115; *p* = 0.005) or COPD (*α*_*LTL*→*T*_ = −0.036; *p* = 0.014) were concordant, they did not survive multiple testing correction. In addition to replicating a previously established correlation between short LTL and increased risk for ischemic heart disease (*β* = −0.024; *p* = 6.6*e*−41) [48], we also found causal evidence for the effect of LTL on ischemic heart disease (*α*_*LTL*→*T*_ = −0.061; *p* = 1.9*e* − 09). Hematological cancer risk negatively correlated with LTL (*β* = −0.015; *p* = 5.8*e*−14), while longer LTL correlated with kidney (*β* = 0.008; *p* = 9.4*e* − 05) and prostate (*β* = 0.029; *p* = 2.1*e* − 23) cancer risk. While we do not have causal estimates for the former, MR confirmed that LTL causally increased risk for kidney (*α*_*LTL*→*T*_ = 0.048; *p* = 8.1*e*−10) and prostate (*α*_*LTL*→*T*_ = 0.089; *p* = 1.0*e*−4) cancers (Figure 2k), aligning with previous findings [49, 17, 20, 50]. This paradox, in which both longer and shorter LTL impact disease risk, was also observed in disorders with an autoimmune component, where shorter LTL is a risk factor for rheumatoid arthritis (*α*_*LTL*→*T*_ = −0.086; *p* = 9.2*e*−5) and Alzheimer’s disease (*α*_*LTL*→*T*_ = −0.037; *p* = 1.3*e*−4), while longer LTL increased risk for systemic lupus erythematosus (*α*_*LTL*→*T*_ = 0.167; *p* = 5.8*e*−5) (Figure 2l-o). Overall, these results highlight the disease-promoting role of both long and short LTL.

### Mediating role of LTL

Analogously to DNA methylation, LTL represents a marker of biological age that can be viewed as a clock integrating a broad range of lifestyle and health parameters [51]. This raises the question whether LTL mediates the relation between complex traits and lifespan. We tested the mediating role of LTL for the relation between 18 non-hematological LTL-modulating traits and lifespan, the latter being affected by LTL at nominal significance (*α*_*LT L*→*T*_ = 0.023; *p* = 0.006). We identified five significant indirect effects (*p*_*indirect*_ *<* 0.05/359), i.e., mediated through LTL (Figure 4a; Table S5). For instance, the negative impact of BMI (*P*_M_ = 7.2%; 95%-CI[3.9%; 10.6%]) or the positive effect of EA (*P*_M_ = 18.8%; 95%-CI[12.3%; 25.7%]) on lifespan were partially mediated by LTL. Given the considerable mediation of AFB (*P*_M_ = 80.8%; 95%-CI[39.4%; 100%]) and ALB (*P*_M_ = 100%; 95%-CI[70.1%; 100%]) on lifespan by LTL, we further investigated these traits through an iterative MVMR approach to build a causal network (Figure 4b; Table S3). Results emphasized the partial mediating role of LTL and EA on the effect of AFB on lifespan.

**Figure 4.**
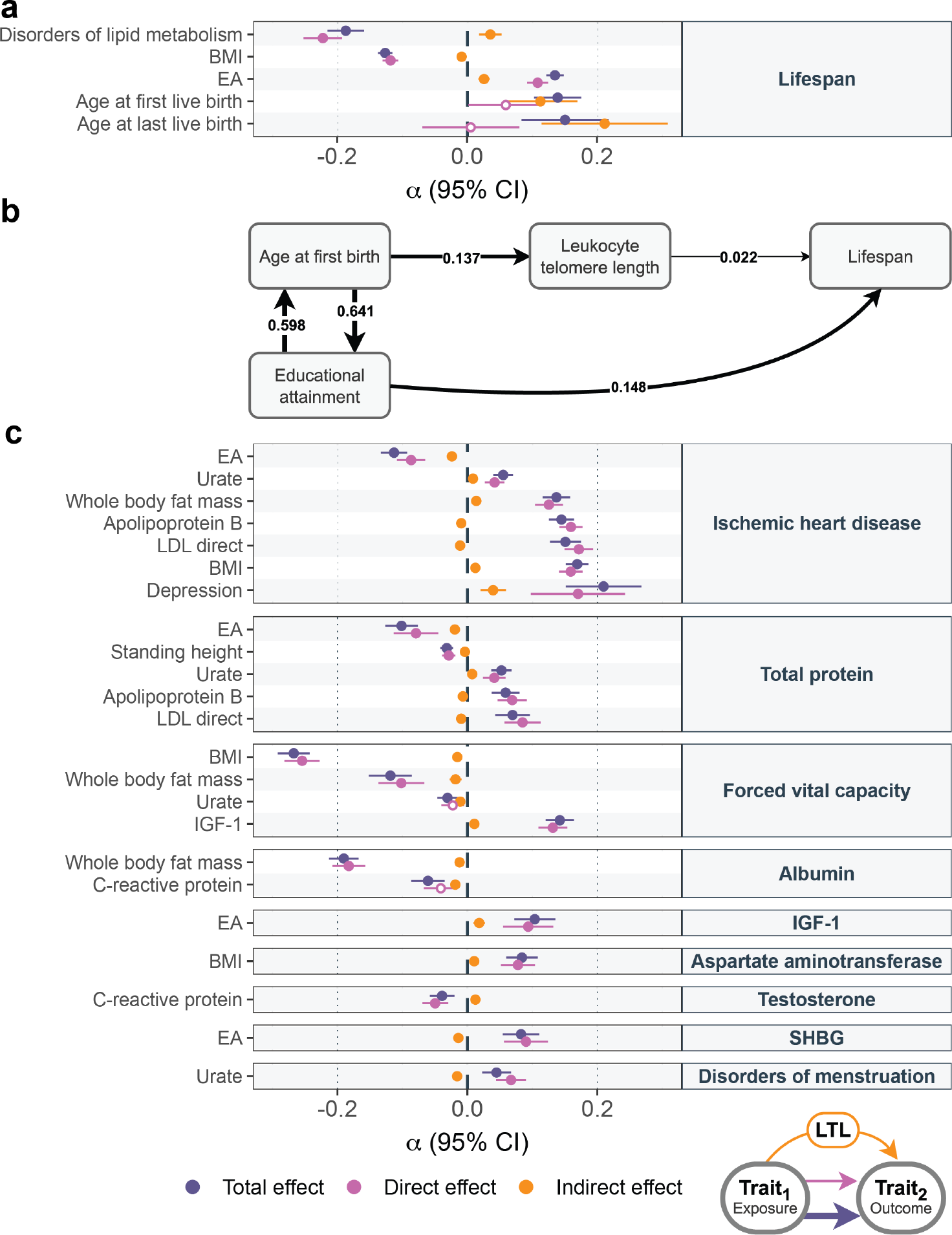
Mediating role of LTL. a) Mediation analysis of 18 LTL-affecting exposures (y-axis; left) on lifespan (y-axis; right) through LTL with effect size estimates and 95% confidence intervals (CI; x-axis) of the total effect (i.e., IVW MR estimate of exposure on outcome; purple), direct effect (i.e., not mediated by LTL; MVMR estimate; pink) and indirect effect (i.e., LTL mediation by product method; orange) as displayed in the scheme in the bottom right corner of the figure. Displayed are relationships with significant (*p* < 0.05/359) total and indirect effects. b) Schematic illustration of the magnitude and direction of nominally significant MVMR effects (*p* < 0.05). Arrow thickness is proportional to the effect size. Significant effects from lifespan to EA are not displayed. c) Mediation analysis of 18 LTL-affecting exposures (y-axis; left) on 19 LTL-affected outcomes (y-axis; right) through LTL. Legend as in (a). EA = educational attainment; LDL = low-density lipoprotein; BMI = body mass index; IGF-1 = insulin-like growth factor 1; SHBG = sex hormone binding globulin; LTL = leukocyte telomere length.

Given that lifestyle factors were found to affect LTL, which in turn influences risk for many diseases, we next used MVMR to assess the LTL mediatory effect for all pairs of 18 LTL modulators and 19 LTL-affected traits. We identified 23 significant (*p*_*indirect*_ *<* 0.05/359) LTL-mediated relationships (Figure 4c; Table S5). Effects on ischemic heart disease, total protein levels, and forced vital capacity were the most frequently mediated by LTL, whereas urate levels, BMI, and EA were the most common exposures. Overall, while we do detect a substantial number of significant mediations through LTL, the average mediation proportion is 5.45%, only accounting for a fraction of these relations.

### Sex differences in LTL biology

Finally, we explored sex differences in LTL biology. We note that overall, there is high concordance between male and female effect estimates, which are highly correlated (*ρ* = 0.92; Figure 5). We sought to identify factors that could explain the stronger age-related decline in LTL observed in males (Figure S1). Given that female reproductive events seemed to impact the rate of telomere shortening in females (Figure 3), we explored whether hormonal effects could account for this difference. Testosterone displayed a stronger association with LTL in males (*p*_diff._ = 4.8*e*−3), while sex hormone binding globulin (SHBG) was associated with LTL in males but not in females (*p*_diff._ = 2.2*e*−18). Correcting LTL for SHBG and testosterone in our sex-stratified regression with age did not abolish the sex difference (*p*_diff._ = 1.3*e*−23). Alternatively, lifestyle factors could account for part of the difference, given that beer consumption (*p*_diff._ = 2.8*e*−6) and smoking status (*p*_diff._ = 3.6*e*−3) showed stronger negative associations with LTL in males, while the association with fruit consumption was more pronounced in females (*p*_diff._ = 9.6*e*−4). After correction for weekly alcohol consumption, alcohol intake frequency, smoking status, as well as beef, vegetable, and fruit consumption, the observed sex difference (*p*_diff._ = 1.0*e*−17) was reduced but not in a significant way (Table S3). Other traits showing a different association with LTL in males versus females include MCH (*p*_diff._ = 8.2*e*−12) and IGF-1 (*p*_diff._ = 5.1*e*−8) which were more strongly associated in males, while hip circumference was more strongly associated in females (*p*_diff._ = 3.3*e*−4). The association with aneurysm was male-specific (*p*_diff._ = 1.5*e*−4), while urea levels (*p*_diff._ = 1.0*e*−4) exhibited a slight negative correlation in females and a positive one in males. Correcting LTL for each of these traits and comparing sex-stratified regression with age did not account for observed differences. Hence, the sex-specific LTL associations identified in our study do not appear to majorly contribute to sex differences in LTL shortening.

**Figure 5.**
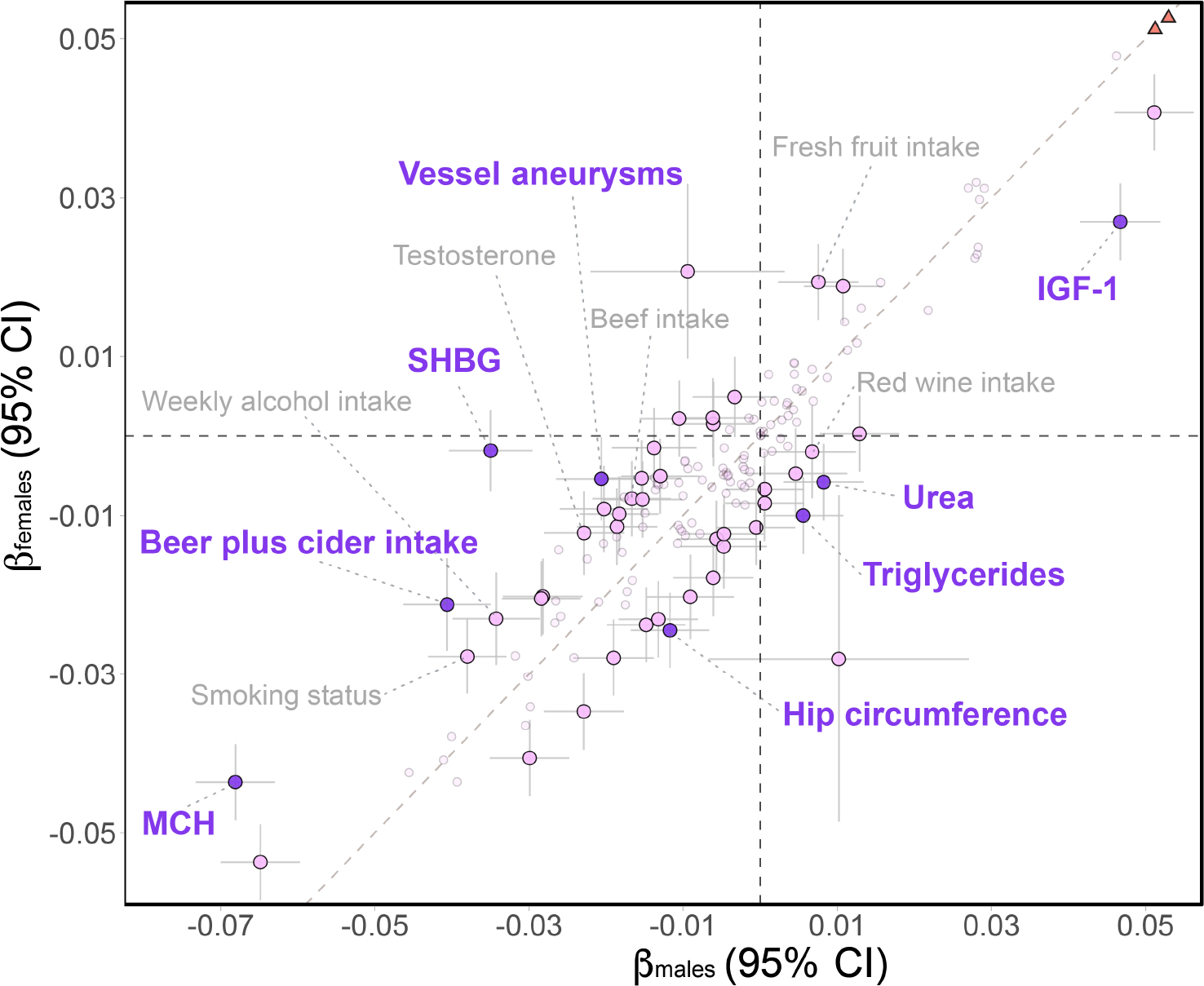
Sex-specific phenome-wide LTL associations. Female-specific (y-axis) against male-specific (x-axis) LTL-trait correlation estimates with 95% confidence intervals. Purple, pink, and smaller gray data points indicate traits with strictly significantly (*p*_diff._ *<* 0.05/141), nominally significantly (*p*_diff._ *<* 0.05), and non-significantly divergent estimates, respectively. Strictly significant traits are labeled in purple, while nominally significant lifestyles or hormonal traits are labeled in gray. Dashed grey and black lines represent the identity line and null effect sizes, respectively. Orange triangles in the top right corner denote non-significant differences in father’s and mother’s age at birth, which are beyond the plotting range (see Table S4).

## Discussion

We comprehensively examined the bidirectional causal relationships between LTL and complex traits, diseases, and lifestyle factors and used MVMR to examine causal effect mediation. Our study reiterates age and sex as major determinants of LTL variability [41, 18] and while hormonal and lifestyle variations showed different associations between males and females, they could not account for the observed sex differences. Still, the fact that lifestyle factors causally affect LTL emphasizes their influence on aging at a physiological and molecular level. Furthermore, we provide evidence for a causal role of abnormal LTL in a broad spectrum of clinically relevant traits, including cancer, autoimmune disorders, lung diseases, and cardiovascular conditions. Lastly, our results show that LTL partially mediates the effect of BMI, EA, and reproductive traits on lifespan.

Others [20, 21] have also used MR to estimate the impact of LTL on 25 traits assessed in our study, all showing concordant effects in our analyses. Notably, we identified a causal effect of LTL on CKD risk that was previously missed, likely due to the reliance of these studies on UKBB summary statistics that were generated based on much lower case counts than the consortia summary statistics used in our study. Earlier research suggested bidirectional causality between telomere attrition and CKD [52]. While we replicated the deleterious impact of shorter LTL on CKD risk, we did not find evidence for a reverse causal effect. Our study also supports the Alzheimer’s disease risk-increasing effect of shorter LTL [53, 54, 55, 56, 57] which was proposed to be driven by promotion of cellular senescence. Paralleling a recent report [58], our results suggest that LTL variation increases the risk for some autoimmune conditions, e.g., risk of systemic lupus erythematosus being increased by longer LTL. Overall, this supports the deleterious role of both long and short telomeres in shaping human health.

Our study also estimated the causal effects of phenotypes on LTL. In line with prior research, alcohol consumption [59], smoking [8, 60], obesity [61], and socioeconomic disadvantages [62] emerged as significant contributors to telomere shortening, underscoring the potential benefits of lifestyle modifications. Some of these factors, such as BMI and EA, were found to exert a small, albeit significant proportion of their impact on longevity through LTL. Surprisingly, the positive influence of serum lipid levels on LTL often attenuated the total effect of lipid-trait relationships. These results are unexpected as high cholesterol levels promote oxidative stress [63], which in turn accelerates LTL shortening [64], warranting further studies to determine mechanisms through which higher lipid levels could favor longer telomeres.

Lifestyle factors modulating LTL, such as alcohol intake, smoking, and dietary habits - which present with strong sex differences and differentially impact LTL in males versus females - represent good candidates to explain the increased rate of LTL shortening in males. Still, we could not demonstrate that these factors were responsible for the differential rate of LTL shortening between sexes. These negative results might be driven by the fact that these results rely on self-reported data that is inherently prone to reporting error [65] and thus represent imperfect proxies of true behaviors, so that further investigations will be required to fully understand sex disparity in LTL shortening. Importance in sex-specific LTL regulators is further highlighted by the finding that delayed AFB and ALB causally associated with longer LTL, an association that was only partially confounded by SES. These results align with the intensified LTL shortening rate we observed after childbirth, which further exacerbated post-menopause. Although we did not observe an association between oestradiol levels - measured only in _∼_49,000 individuals - and LTL, we hypothesize that hormonal shifts following pregnancy and menopause could accelerate LTL shortening [66]. An alternative explanation is that LTL shortening is driven by the stress imposed by such events on the body. While further research is required to test these hypotheses, our results highlight the prominent role of life history events in LTL shortening rates. Overall, this emphasizes how hormonal and lifestyle factors can influence LTL, which in turn impacts global health and disease risk through complex networks.

Our study is subject to several limitations. First, the use of cross-sectional bulk LTL limits our capacity to analyze individual telomere shortening rates, which might be a critical factor in disease prediction [51]. Second, although LTL and telomere length in other tissues are correlated [67], this proxy might miss more subtle and tissue-specific relations between telomere length and the phenome. Furthermore, we cannot exclude that the tissue in which telomere length was measured (i.e., leukocytes) biases the observed associations with hematological traits, even if accounting for blood cell type traits did not seem to affect association estimates between LTL and other traits. Third, our study focused on White-British ancestry, meaning the results may not necessarily apply to other ethnicities. In the future, single-cell telomere length measured at chromosomal resolution through long-read sequencing approaches across a wide variety of tissues, time points, and ancestral groups should provide a more refined view of telomere dynamics. Fourth, MR presents with inherent restrictions, notably susceptibility to horizontal pleiotropy violations, especially given the considerable heterogeneity across our IVs. In that optic, we used a broad range of sensitivity analyses and focused on results robust across these various methods. Another limitation of MR is that detection power is bound by the number of available IVs, so that our power to detect causal relations between traits and LTL is variable across phenotypes and might be lower or larger than for the reverse LTL on trait relation, depending on whether the trait has less or more IVs than LTL, respectively. Finally, MR does not account for dynamic spatiotemporal changes in LTL that occur over lifetime and/or in the context of some diseases such as cancer.

## Conclusion

Through usage of univariable and multivariable bidirectional Mendelian randomization, we identify a complex net-work of causal relations wherein both exogenous and endogenous environmental factors modulate LTL, which in turn influences the risk for numerous diseases and mediates the impact of some of these traits on lifespan. Still, based on currently available data, its mediatory role between unfavorable lifestyle and disease is estimated to be modest, and further research is needed to explore the relation between LTL and other aging biomarkers, such as DNA methylation, in order to understand its clinical value as a proxy of biological age.

## Data Availability

UK Biobank data is accessible to registered users, while GWAS data is freely available, as detailed in Supplementary Table 2.

https://pan.ukbb.broadinstitute.org

http://www.nealelab.is/uk-biobank/

## Data and code Availability

All data used in this study are publicly available, as described in the methods. Code to reproduce analyses per-formed in this study is available at https://github.com/cChiiper/UNIL_SGG_MR_LTL

## Acknowledgments

This research has been conducted using the UK Biobank Resource under Application Number 16389. We thank all UK biobank participants for sharing their data. Computations were performed on the JURA and Urblauna servers from the University of Lausanne. The study was funded by the Swiss National Science Foundation (310030 189147, ZK) and the Department of Computational Biology of the University of Lausanne (ZK).

## Author contributions

SM, CA, and ZK conceived the study; SM carried out the analyses with contributions from MCS; ZK supervised statistical analyses; SM generated the figures; SM and CA drafted the manuscript and ZK made critical revisions; All authors read, approved, and provided feedback on the final manuscript.

## Ethics declarations

The authors declare that they have no competing interests.

## Supplementary information

### Supplementary Information

Supplementary Note and Figures 1-9.

### Supplementary Tables

Supplementary Tables 1-5.

