## Supplementary Information for "Breaking down causes, consequences, and mediating effects of age-related telomere shortening on human health"

### Supplementary Note

#### Composite trait definition

Composite traits are defined as follows:

- *Age at first and last live birth*: Age at first live birth (#2754) and age at last live birth (#2764) were completed with age of primiparous women at birth of child (#3872) as females with single pregnancy are not factored in the two first fields.
- *Average hand grip*: Average of left (#46) and right (#47) hand grip strength.
- *Cholesterol medication*: Patients under cholesterol-lowering medication were selected based on UKBB data fields #6177 (males) and #6153 (females).
- *Educational attainment*: We resitricted age at full-time education completion (#845) between 14 and 19 years. The 413 participants with an age under 14 years were set to 14, while the 20,669 with an age over 19 were set to 19. The 102,884 participants with a College or University degree (#6138) were attributed an education completion age of 19.
- *Mother's and father's age at birth*: Age (#21003) was subtracted from mother's age (#1845) and father's age (#2946), respectively, for those with living parents.
- *Proxied Lifespan*: For deceased participants, scaled age at death (#40007) in a sex-specific fashion; For living participants, average between scaled mother's (#3526) and father's (#1807) age at death if both are available, otherwise age at death of the available parent. Scaling was performed in a sex-stratified manner to account for the differences in life expectancy between sexes.
- *Reproductive lifespan*: Age at menarche (#2714) was subtracted from age at menopause (#3581).
- *Waist-to-hip ratio*: Waist cirumference (#48) over hip circumference (#49).
- *Waist-to-hip ratio adjusted for BMI*: Waist-to-hip ratio corrected for body mass index (BMI) (#21001) in a sex-stratified manner.
- *Weekly alcohol intake in units*: The average weekly consumption of each type of alcohol, such as beer plus cider (#1588), champagne plus white wine (#1578), fortified wine (#1608), red wine (#1568), spirits (#1598), and other alcoholic drinks (#5364), was summed together. Participants with an average of multiple instances were rounded to one decimal. Cases, where all data fields were not answered, were set as missing. Participants drinking alcohol with missing information in some data fields were completed with the rounded average alcohol uptake of each data field (i.e., #1588: 3.0 units, #1578: 2.7 units, #1608: 0.2 units, #1568: 3.9 units, #1598: 1.8 units, #5364: 0 units)

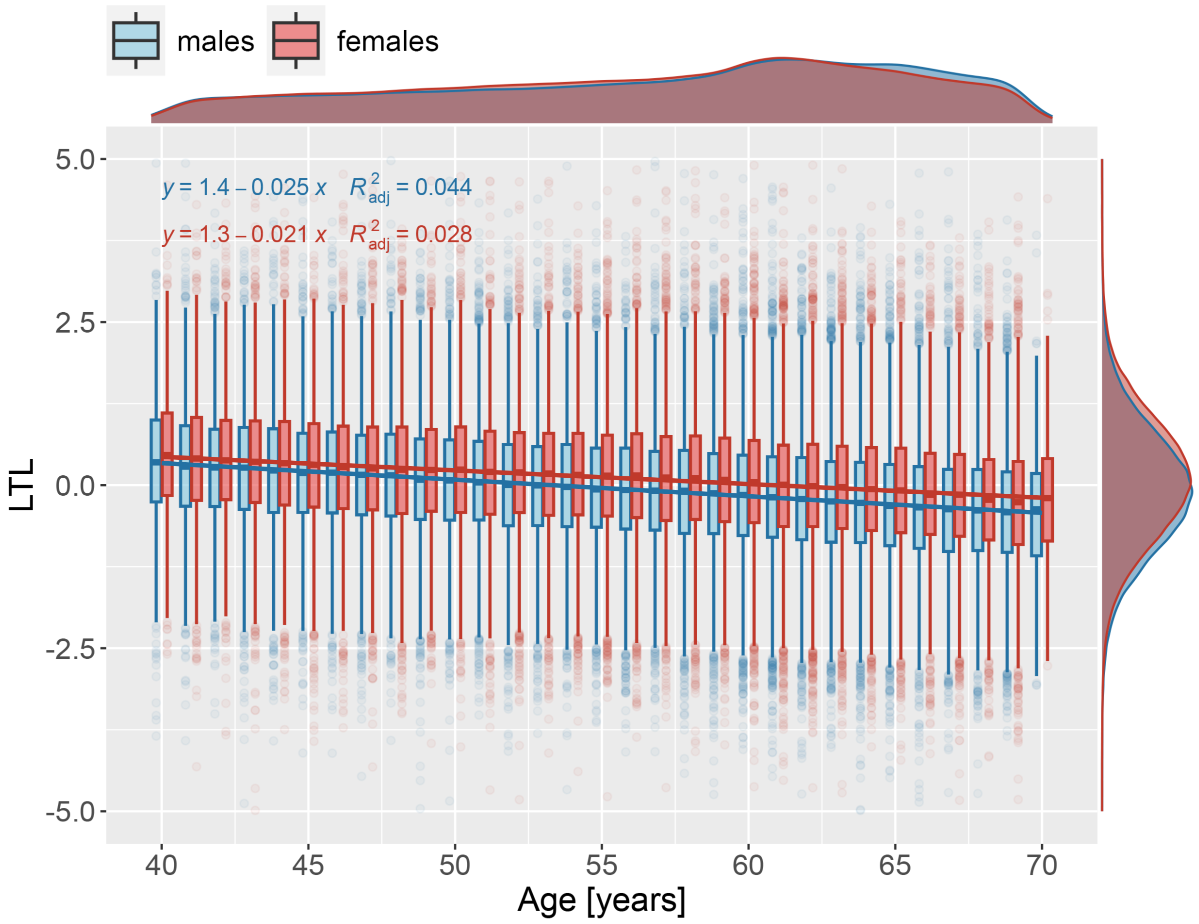


**Supplemental Figure 1. Linear regression of LTL against age, stratified by sex**

Boxplots of Z-adjusted leukocyte telomere length (LTL; y-axis; distribution in margin) in function of participant's age (x-axis; distribution in margin), stratified by sex. Extreme outliers (mean $\pm$5 SD; $N = 107$), are not shown. Regression equations with adjusted r-squared for males (blue) and females (red) are indicated in the upper left corner.

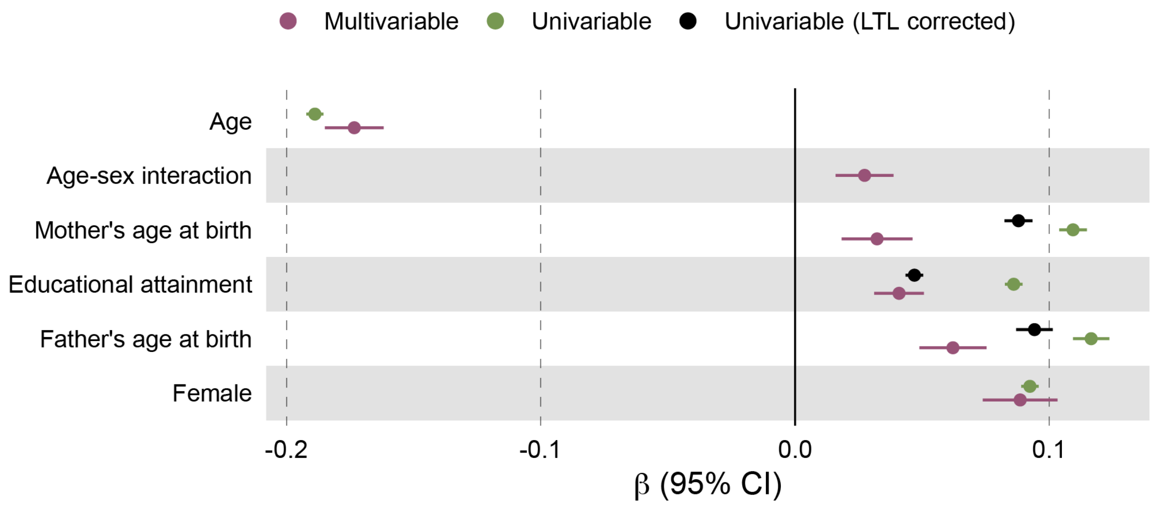


**Supplemental Figure 2. Decomposition of the effects of mother's and father's age at birth on LTL**

Linear regression estimates (x-axis) with 95% confidence intervals (CI) of traits (y-axis) on LTL from multivariable (i.e., all traits jointly assessed in the same model; pink), univariable with unadjusted LTL (green), and univariable with covariate-corrected LTL (black) models (see Table S3).

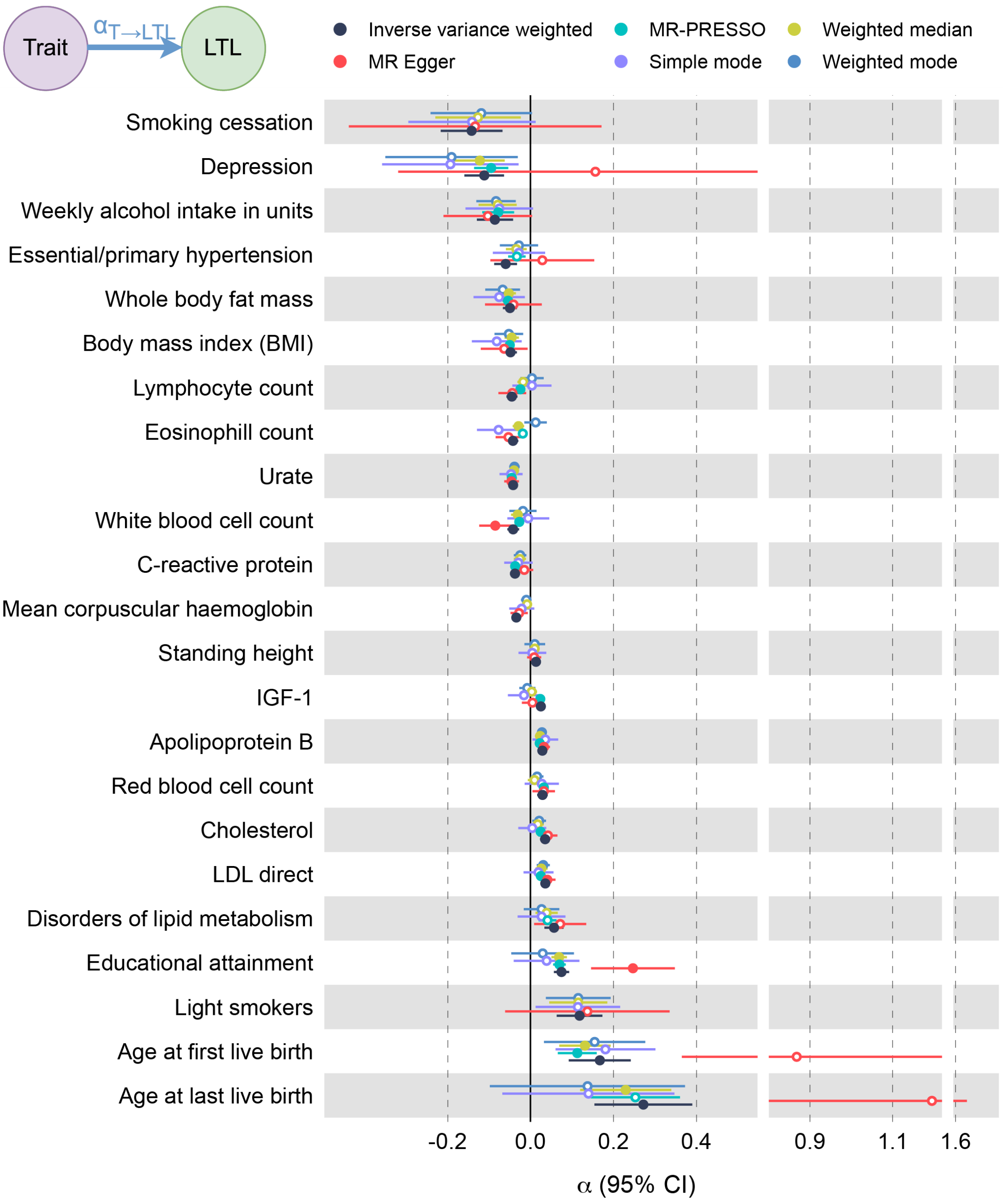


**Supplemental Figure 3. MR estimates of trait on LTL effects across different methods**

Mendelian randomization (MR) estimates (x-axis; broken) with 95% confidence intervals (CI) for traits (y-axis) with strictly significant ($p < 0.05/141$) inverse variance weighted MR (IVW) effect on LTL. Estimates for 6 MR methods are shown: IVW (black), MR-PRESSO (teal), weighted median (green), MR Egger (red), simple mode (purple), and weighted mode (blue). Strictly significant effects are shown as full circles; otherwise as empty circles. MR-PRESSO are missing for i) light smokers and smoking cessation due to the absence of identified outliers and ii) standing height and mean corpuscular hemoglobin due to convergence issues. IGF-1 = insulin-like growth factor 1; LDL = low-density lipoprotein (see Table S4).

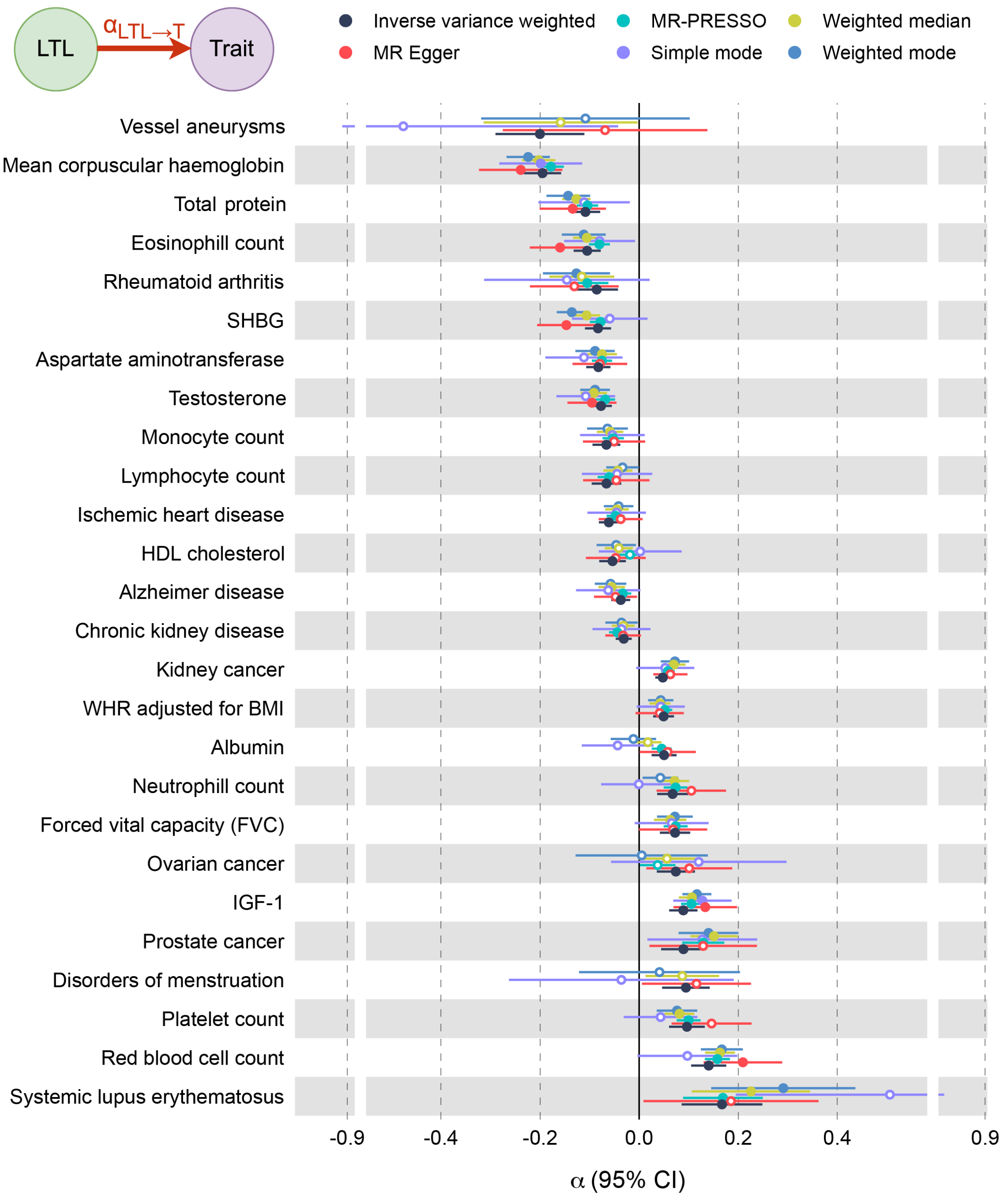


**Supplemental Figure 4. MR estimates of LTL on traits across different methods**

Mendelian randomization (MR) estimates (x-axis; broken) with 95% confidence intervals (CI) for traits strictly significantly ($p < 0.05/141$) affected by LTL (y-axis) based on the inverse variance weighted MR (IVW) effect. Estimates for 6 MR methods are shown: IVW (black), MR-PRESSO (teal), weighted median (green), MR Egger (red), simple mode (purple), and weighted mode (blue). Strictly significant effects are shown as full circles; otherwise as empty circles. MR-PRESSO estimates for vessel aneurysms and disorders of menstruation are missing due to the absence of identified outliers. SHBG = sex hormone binding globulin; HDL = high-density lipoprotein; WHR = waist-to-hip ratio; BMI = body mass index; IGF-1 = insulin-like growth factor 1 (see Table S4).

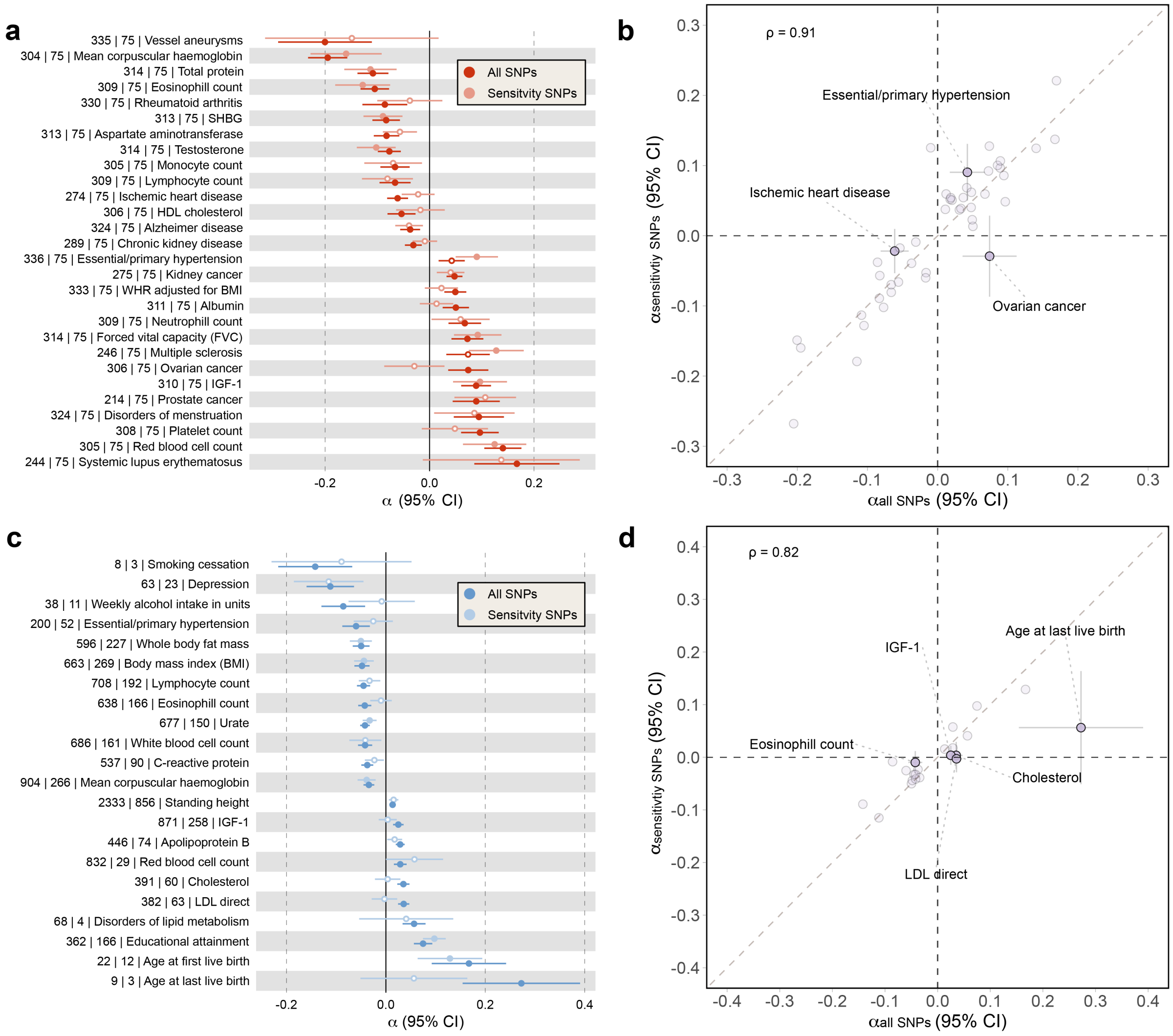


**Supplemental Figure 5. Stringent Steiger pleiotropy-sensitivity analysis**

a) Mendelian randomization (MR) estimates (x-axis) with 95% confidence intervals (CI) for strictly significantly $(p < 0.05/141)$ LTL on trait (y-axis) effects based on inverse variance weighted MR (IVW; red) or stringent IVW (salmon). Sensitivity SNPs were selected based on Steiger filtering $(Z \leq -1.96)$ for 152 traits with available summary statistics. Numbers preceding trait descriptions (y-axis labels) indicate the number of SNPs for total IVW estimates and stringent IVW estimates. Strictly significant $(p < 0.05/141)$ effects are shown as full circles; otherwise as empty circles. b) Stringent IVW estimates for LTL on traits (y-axis) plotted against total IVW estimates of LTL on traits (x-axis) with 95% CIs. Purple and grayed data points indicate traits with nominally significant $(p_{\text{diff.}} < 0.05)$ and non-significantly divergent estimates, respectively. Traits with $p_{\text{diff.}} < 0.05$ are labeled. Dashed grey and black lines represent the identity line and null effect sizes, respectively. Pearson correlation displayed in the top left. c) Mendelian randomization (MR) estimates (x-axis) for traits with 95% CI for strictly significant $(p < 0.05/141)$ trait on LTL IVW MR (blue) or stringent IVW (light blue). Rest of the legend as in a). d) Stringent IVW estimates for trait on LTL (y-axis) plotted against total IVW estimates of traits on LTL (x-axis) with 95% CIs. Rest of the legend as in b). SHBG = sex hormone binding globulin; HDL = high-density lipoprotein; WHR = waist-to-hip ratio; BMI = body mass index; IGF-1 = insulin-like growth factor 1; LDL = low-density lipoprotein (see Table S4).

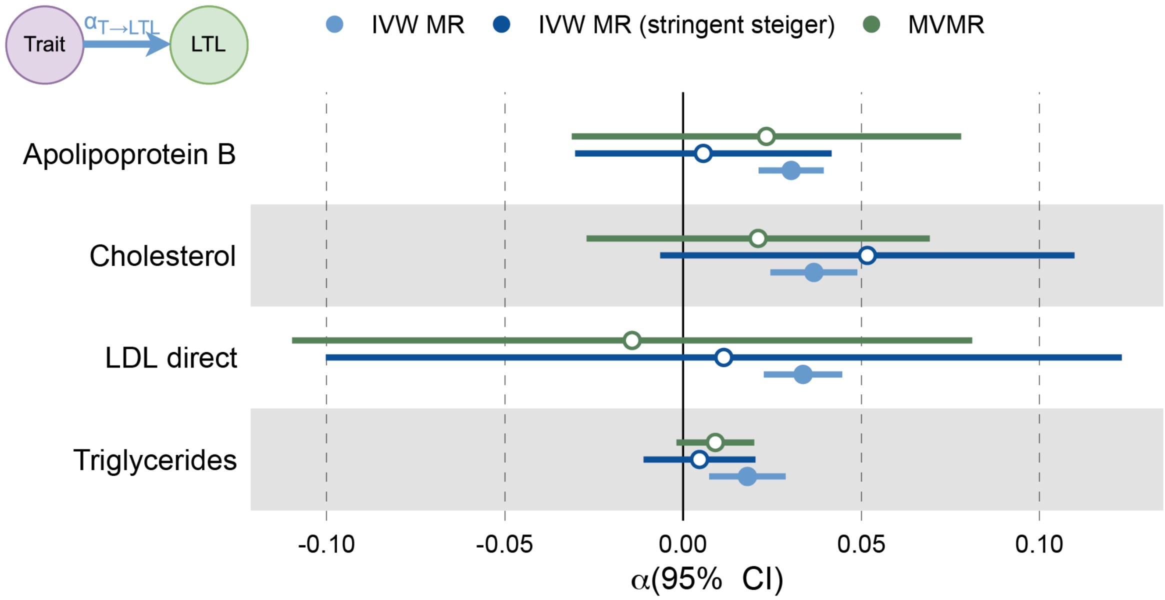


**Supplemental Figure 6. MVMR effect estimation of traits on LTL**

Mendelian randomization (MR) estimates (x-axis) with 95% confidence intervals (CI) of lipid traits (y-axis) on LTL. Estimates obtained from inverse variance weighted (IVW; blue), stringent IVW (Steiger filtering between all exposures (see Methods); dark blue), and Multivariable Mendelian Randomization (MVMR; green) are shown. Strictly significant ($p < 0.05/141$) effects are shown as full circles; otherwise as empty circles. LDL = low-density lipoprotein (see Table S3).

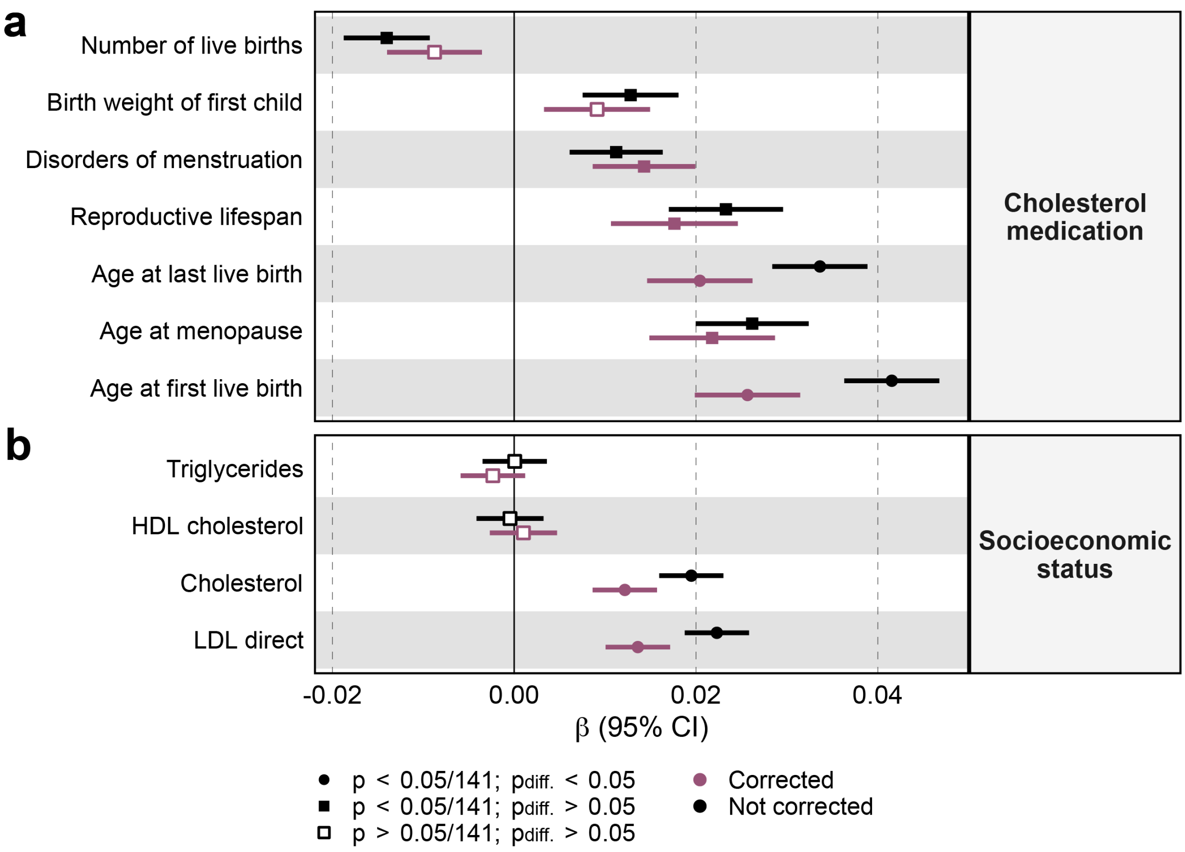


**Supplemental Figure 7. LTL associations adjusted for potential confounding variables**

Linear regression estimates (x-axis) with 95% confidence intervals (CI) of a) female reproductive traits or b) serum lipid levels (y-axis, left) on LTL, adjusted (pink) or not (black) for potential confounders (y-axis, right), i.e., a) socio-economic status or b) cholesterol medication. Significantly different effects $p_{\text{diff.}} < 0.05$ are shown as circles, otherwise squares. Strictly significant $(p < 0.05/141)$ effects are shown as full symbols; otherwise as empty symbols. LDL = low-density lipoprotein; HDL = high-density lipoprotein (see Table S4).

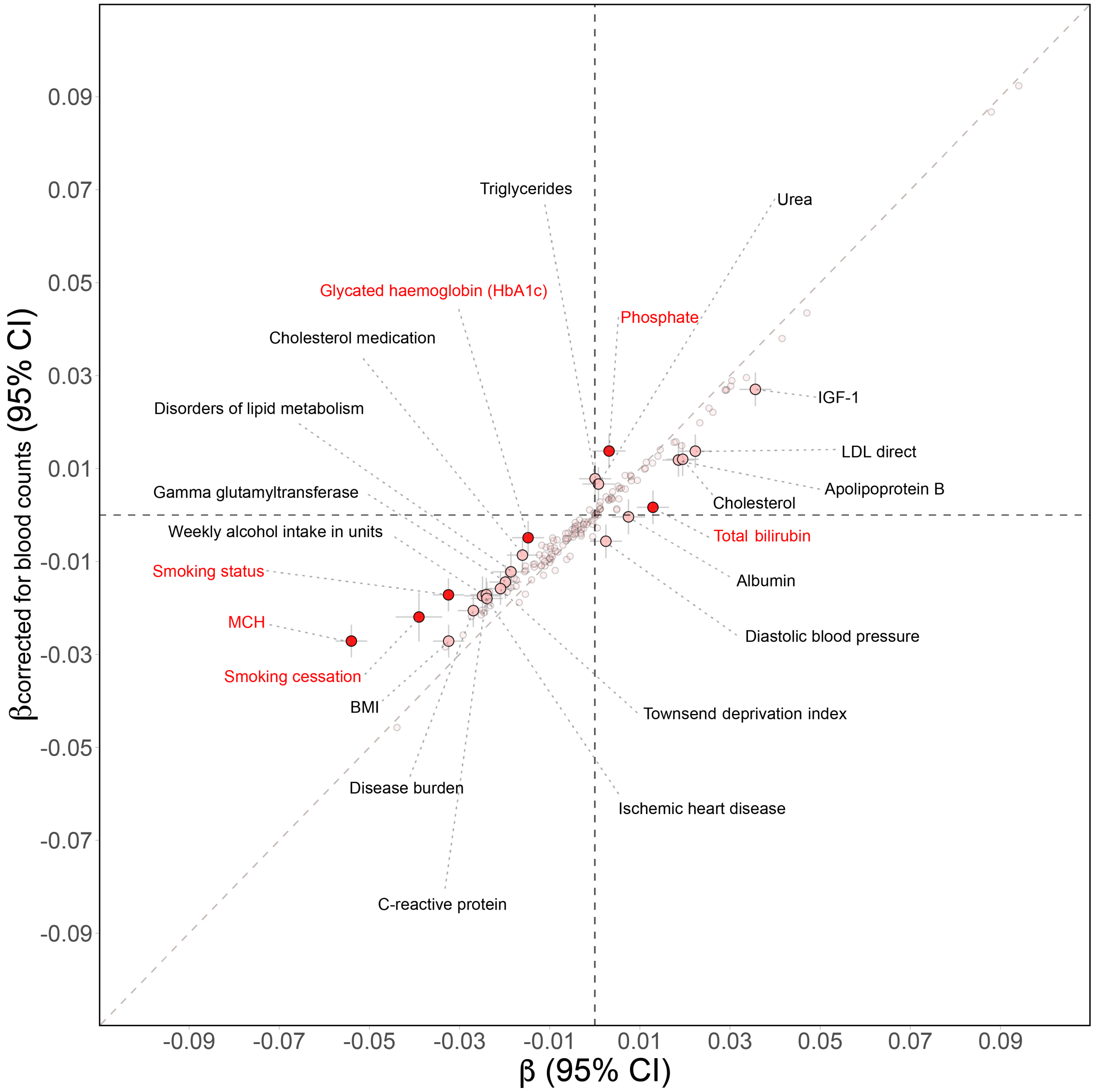


**Supplemental Figure 8. Linear regression estimates adjusted for blood counts**

Observational correlation between LTL and 158 complex traits adjusted for standard covariates-only (x-axis) versus standard covariates plus lymphocyte, white blood cell, red blood cell, platelet, monocyte, eosinophil, neutrophil, and reticulocyte count (y-axis) with 95% confidence intervals (CI). Red, salmon, and smaller gray data points indicate traits with strictly significantly ($p_{\text{diff.}} < 0.05/141$), nominally significantly ($p_{\text{diff.}} < 0.05$), and non-significantly divergent estimates, respectively. Traits with $p_{\text{diff.}} < 0.05$ are labeled. Dashed grey and black lines represent the identity line and null effect sizes, respectively. MCH = mean corpuscular hemoglobin; BMI = body mass index (see Table S4).

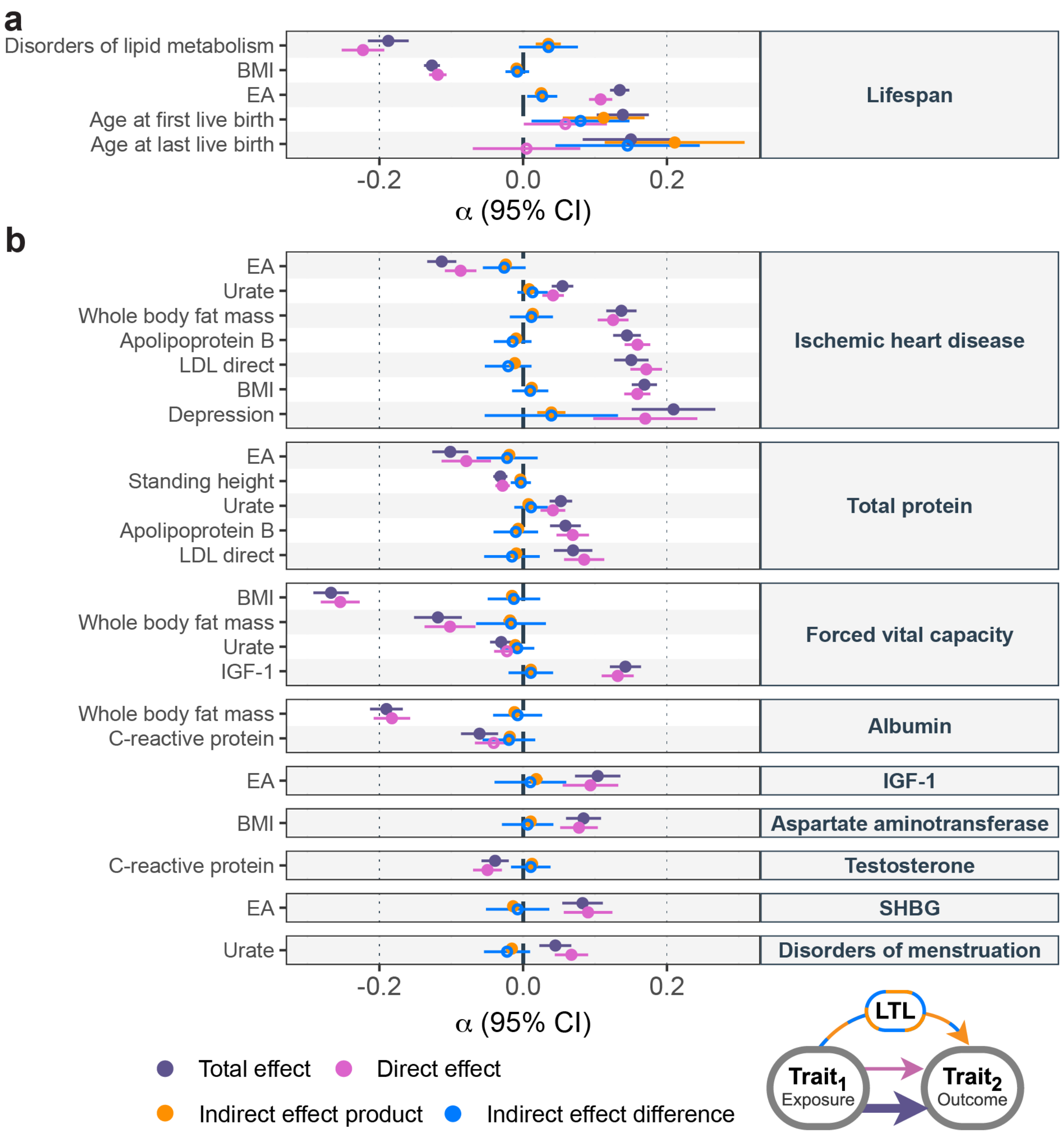


**Supplemental Figure 9. Mediating role of LTL on complex pair trait relations**

a) Mediation analysis of 18 LTL-affecting exposures (y-axis; left) on lifespan (y-axis; right) through LTL with effect size estimates and 95% confidence intervals (CI; x-axis) of the total effect (i.e., IVW MR estimate of exposure on outcome; purple), direct effect (i.e., not mediated by LTL; MVMR estimate; pink) and indirect effect (i.e., LTL mediation by product method; orange and by difference method; blue) as displayed in the scheme in the bottom right corner of the figure. Displayed are relationships assessed on Figure 4. b) Mediation analysis of 18 LTL-affecting exposures (y-axis; left) on 19 LTL-affected outcomes (y-axis; right) through LTL. Legend as in (a). EA = educational attainment; LDL = low-density lipoprotein; BMI = body mass index; IGF-1 = insulin-like growth factor 1; SHBG = sex hormone binding globulin; LTL = leukocyte telomere length (see Table S5).
